# Patient Reported Outcomes of Medical Cannabis for Managing Pain in Charcot-Marie-Tooth Disease

**DOI:** 10.1101/2022.08.09.22278591

**Authors:** Priscilla C. Canals, Alexia G. Aguilar, Gregory T. Carter, C. Miyabe Shields, Andrew Westerkamp, Meg D’Elia, Joy Aldrich, Robert N. Moore, Allison T. Moore, Brian J. Piper

**Affiliations:** Department of Medical Education, Geisinger Commonwealth School of Medicine, Scranton, PA, United States of America; Saint Luke’s Rehabilitation Institute, United States of America; Real Isolates LLC, Beverly Massachusetts, United States of America; CeresMed, South Burlington, Vermont, United States of America; Hereditary Neuropathy Foundation, New York, NY, United States of America; Center for Pharmacy Innovation and Outcomes, Precision Health Center, Forty Fort, PA, United States of America

**Keywords:** hereditary motor and sensory neuropathy, neuropathic pain, marijuana

## Abstract

**Background and aims:** Chronic pain is a major part of the disease burden in Charcot-Marie-Tooth (CMT) disease. Current pharmacotherapies to manage symptoms of CMT disease, particularly pain, are inadequate. This exploratory study examined the patient reported efficacy of medical cannabis among CMT patients.

**Methods:** Participants (N = 56; 71.4% female; Age = 48.9, SD = 14.6; 48.5% CMT1) were recruited though the Hereditary Neuropathy Foundation’s Global Registry for Inherited Neuropathies. The online survey contained 52 multiple choice questions about demographics, medical cannabis use, symptomology, efficacy, and adverse effects.

**Results:** When asked about how much relief they experience from using cannabis as a method of symptom relief, respondents reported an average of 69.6% (SEM ± 2.6). Women were more likely to report experiencing pain than men (*p* < .05). Participants who perceived support from their providers were more likely to inform them of their cannabis use (*p* < .05).

**Interpretation:** Patients reported that cannabis was effective to manage symptoms. More prospective and controlled research needs to be conducted to better serve and optimize the potential use of cannabis to treat CMT.

**Graphical Abstract:** Patient Reported Outcomes of Medical Cannabis for Managing Pain in Patients with Charcot-Marie-Tooth Disease

Priscilla C. Canals, Alexia G. Aguilar, Gregory T. Carter, Marion McNabb, Andrew M. Westerkamp, Miyabe Shields, Meg D’Elia, Joy Aldrich, Robert N. Moore, Allison T. Moore, Brian J. Piper^*^

- There is no prior research of medical cannabis experiences among Charcot-Marie-Tooth (CMT) patients.
- CMT patients (N=56) were recruited though the Hereditary Neuropathy Foundation’s Global Registry for Inherited Neuropathies and completed an online survey.
- Symptom relief from using cannabis was moderately-high (70% ± 3).
- CMT patients that received support from their providers were significantly more likely to inform them of their cannabis use.
- These descriptive results should be verified using prospective and randomized controlled trials.

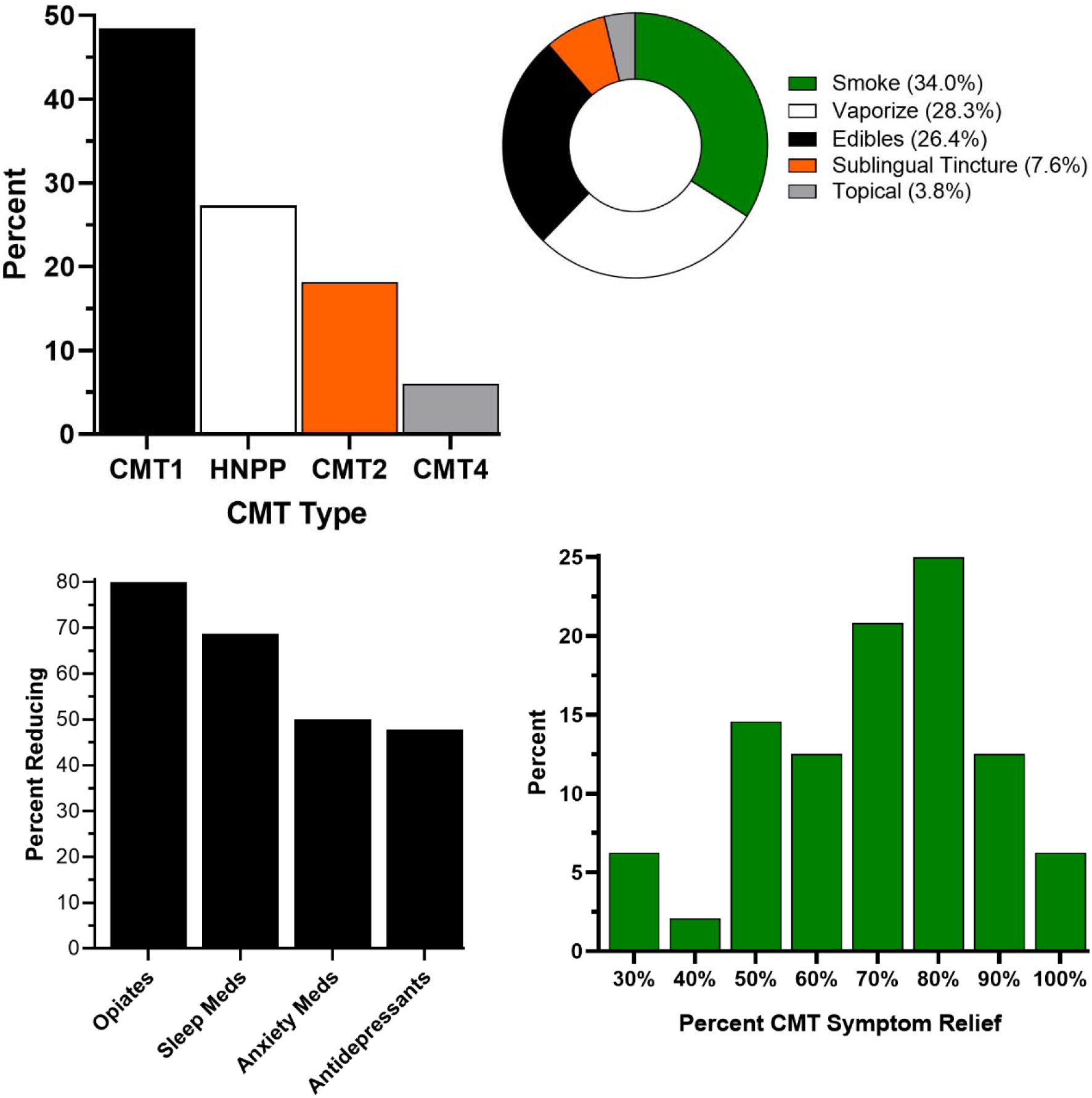

## Introduction

Charcot-Marie-Tooth (CMT) disease is the most common genetic neuropathy affecting 1 in 2,500 people [1]. Although the motor symptoms may attract the most attention, a large survey (N = 407) revealed that over three-quarters of patients with CMT experienced pain and over four-fifths numbness [2]. Among genetically diagnosed patients with CMT1A, 29% had neuropathic pain. Allodynia was the most specific neuropathic pain symptom [3]. Complications of CMT include sleep apnea, restless legs syndrome, daytime sleepiness, and impaired sleep [4].

Although there is no known cure for this slowly progressive disease, there are treatments such as physical and occupational therapy, orthotics, and pharmacotherapies to manage symptoms. The medications used for pain management are varied and included aspirin, nonsteroidal anti-inflammatory medications, acetaminophen, tricyclic antidepressants, anticonvulsants, and opioids [5].

The National Academy of Sciences reported that there was conclusive evidence indicating that cannabis was effective for chronic pain among adults [6]. There was also moderate evidence that cannabis was effective for improving short-term sleep outcomes in individuals with sleep disturbance associated with obstructive sleep apnea syndrome or chronic pain [6]. Clinical-practice guidelines are now emerging recommending that chronic pain patients can be treated with medical cannabis [7]. Surveys of cannabis dispensary members have identified a harm-reducing substitution effect where they reduce or stop using other medications, especially opioids [8], after starting cannabis. There is currently no published information about medical cannabis that is specific to CMT. The objective of this exploratory study was to obtain CMT patient’s perspectives regarding the benefits and risks of medical cannabis.

## Materials & Methods

### Participants

Participants (N = 56, 71.4% female, Age = 48.9, SEM=2.0, Min = 22, Max = 87) were recruited for an online survey. Among the subset with a known CMT type (N = 33), this was 48.5% CMT1, 27.3% Hereditary neuropathy with liability to pressure palsies (HNPP), 18.2% CMT2, and 6.1% CMT4. Among those that reported using a motility device (N=24), this included cane (25.0%), braces (20.8%), walker (16.7%), and scooter (12.5%).

### Procedures

The survey used for this study included a subset of the items used previously [8] but also many questions were tailored for the CMT population. The survey contained 52 multiple choice and short-answer items (Supplemental Appendix) and took about ten-minutes to complete. The instrument gathered information on the utility, strain preference (e.g. tetrohydrocannabinol (THC) vs cannabidiol (CBD)) and adverse effects of medical cannabis. Survey links were posted on the Hereditary Neuropathy Foundation website, a US based patient advocacy organization [9], and sent to all members. Inclusion criteria were those affected by CMT or inherited neuropathy who were over the age of 18, and who used medical cannabis to cope. Participation was voluntarily and anonymous. The study was IRB approved as exempt by Advarra.

### Data-analysis

Data was imported into IMB SPSS, version 26 to conduct chi-square tests. Figures were constructed with GraphPad Prism, version 8. The variability was reported as the Standard Error of the Mean (SEM). A *p* < .05 was considered statistically significant.

## Results

Issues related to muscle weakness and atrophy were common in this sample. About three-quarters of respondents reporting weak ankles (72.2%), about one-third with muscle atrophy (36.1%), tremors (33.3%); a quarter had poor or absent reflexes (27.8%), clumsiness (25.0%), coordination or balancing problems (25.0%), restless leg syndrome (25.0%), arm or leg weakness (22.2%), difficulty with physical activity (22.2%), rigid muscles (22.2%); and a fifth had hammer or curled toes (19.4%), spasticity (19.4%), and weakness (19.4%). Sensory concerns included temperature sensitivity (27.8%), neuropathic pain (22.2%), and numbness (22.2%).

Preferred methods of cannabis consumption were smoking (34.0%), vaporize (28.3%), edibles (26.4%), sublingual tinctures (7.6%), and topical (3.8%). Preferred strains were indica dominant (22.2%), 50/50 THC/CBD (20.4%), high CBD (16.7%), high THC strains (9.3%), sativa dominant strains (5.6%), and hybrid strains (3.7%). One-third (33.9%) possessed a medical cannabis certificate.

When asked if they experienced pain associated with CMT, 90.9% of participants answered ‘yes’. All (100%) females but only 72.7% of males reported pain associated with CMT (chi-square *p* < .05). In response to: ‘How effective is medical cannabis in alleviating your symptoms with CMT?’ respondents provided the percent of relief they experienced from medical cannabis (0 to 100% in 10% increments). The most frequently selected option was 80% relief. The vast majority (91.7%) indicated cannabis provided at least 50% relief (Figure 1A).

**Figure 1.**
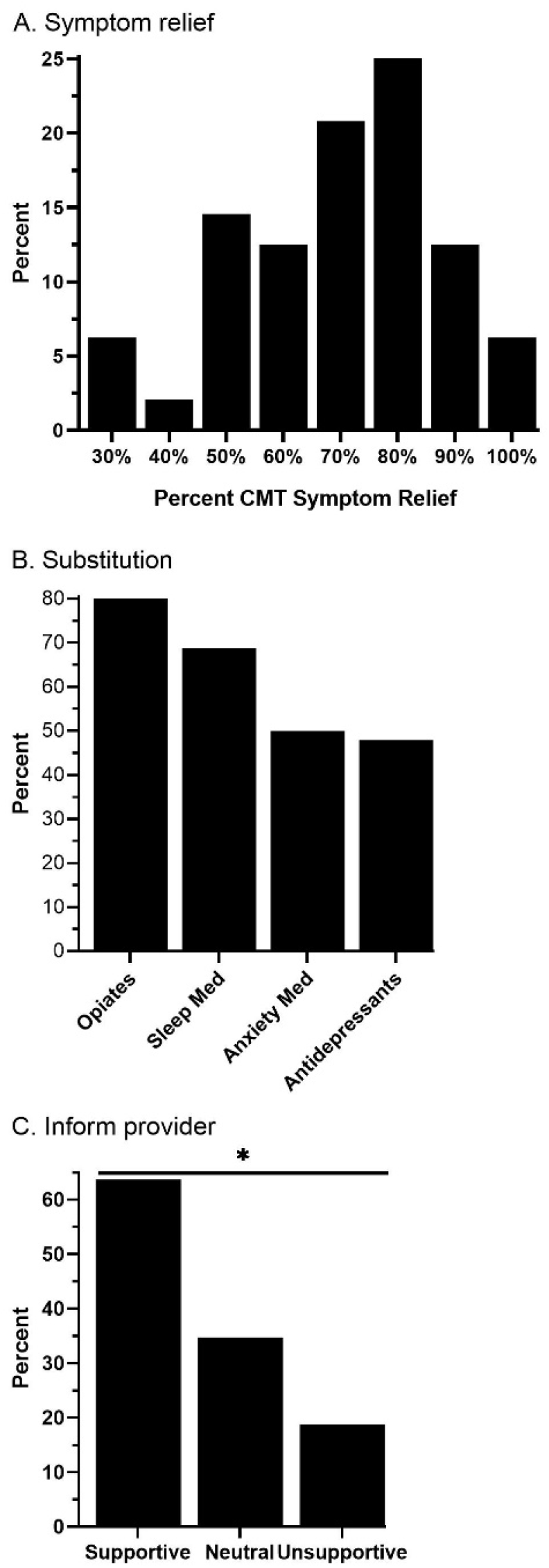
Charcot-Marie-Tooth patients’ (N = 56) evaluations of medical cannabis. Percent symptom relief (mean = 69.6, SEM= 2.6) from medical cannabis (A). Percent reporting a reduction in medication use after using medical cannabis (B). Percent that informed health-care providers of cannabis use based on perceived support (C, \**p* < .05 versus unsupportive).

Participants were asked if they noticed a change in their use of other medications after they began using medical cannabis. Among the subset that used each drug class, four-fifths (80.0%) reported using less opiates, two-thirds (68.8%) with less sleep medication, and half (50.0%) with less anxiety medications or less antidepressants (47.8%, Figure 1B).

Physicians’ attitudes regarding patient medical cannabis use were examined. For those who reported receiving a supportive response from providers, three-fifths (63.6%) indicated that they would inform them of their medical cannabis use. For those who reported unsupportive responses from providers for medical cannabis use, less than one-fifth (18.8%) said they would inform providers of their cannabis use (*p* < .05, Figure 1C,).

Participants reported whether they experienced negative side effects due to cannabis use (23.5% do) and if they planned on halting their usage (2.0% answered ‘yes’ and 9.8% ‘unsure’). The preponderance who did (91.7%) as well as most who did not (87.2%) have negative side effects did not have plans to stop consuming medical cannabis.

## Discussion

This is the first study examining patients with CMT perspectives regarding MC. Patients with CMT reported that medical cannabis provided marked symptom relief. Based on the substantial (70%) improvement in CMT symptoms, cannabis may be a promising therapeutic option if these findings are verified with prospective or randomized controlled trials. These findings extend upon prior research with cannabinoids as complementary and alternative therapeutic agents for other neurological conditions [6, 10]. The results for multiple-sclerosis appear to be formulation dependent [11, 12]. The American Academy of Neurology reports that there is high quality evidence indicating that oral cannabis extract is efficacious in relieving symptoms of spasticity and pain in patients with multiple sclerosis. There were also concerns about adverse effects as well as the use of non-standardized cannabis extracts [13].

Perceived provider attitudes may adversely affect patients’ likelihood to inform providers of their medical cannabis use. There have been some, mostly regionally focused, efforts to characterize providers attitudes towards medical cannabis [14-16], particularly among medical oncologists [17-18]. Medical cannabis is a controversial topic and the likelihood of an open-dialogue between patients and providers likely varies based on provider specialty, region of the country, and employer policies [8, 17, 19]. Cannabis was viewed by these patients as a therapeutic drug and, as such, it is crucial that they are able to inform their medical providers of their use.

This report also determined that four out of five CMT patients reported decreasing their use of opiates after starting medical cannabis. Similarly, two-thirds (68.8%) reported less sleep medication, and half (50.0%) with less anxiety medications or less antidepressants (47.8%). The identified substitution effect corroborates prior investigations including among New England dispensary patients primarily with chronic pain [8], Prescription Drug Monitor Program records in New Mexico [20] and Medicare and Medicaid prescription records [21, 22]. However, recent ecological research has challenged whether this substitution effect meaningfully impacts opioid overdoses [23, 24]. Regardless, there would appear to be some harm reduction impact of cannabis by allowing patients to take less opioids. However, further attention is needed if those that use less prescription medications (e.g. antidepressants) are also not informing their health care providers.

Also noteworthy was that only one-third of participants in this study were certified for medical cannabis. The demarcation between medical and recreational cannabis use is likely complex [8] as some “medical users” may use it for recreational purposes and some “recreational users” may be self-treating for therapeutic purposes. The cost of medical cannabis is non-trivial and is not covered by insurance.

There are a number of notable limitations to this study. Inherent to survey studies is respondent bias. All participants (N=56) in this study use cannabis. Given the branched design of the survey, there were multiple questions where fewer than twenty respondents provided information. This would bias the dataset toward a positive cannabis experience as those who are more inclined to answer the survey generally have more favorable experiences with medical cannabis. Although the sample size (N = 56) was only moderate, CMT is a relatively rare disease [1] making these findings particularly novel relative to the absence, to our knowledge, of prior investigations. Cannabis not being used in a controlled setting (dosage, composition, methods of delivery, etc.) was also a limitation. Cannabis can be administered in many ways, whether it is inhaled through vapor or ingested in food or drinks and each method has a different rate of absorption. Although some participants reported a decrease in their pain, there is no indication as to how much cannabis they consumed in order for them to see that decrease. It is possible that those who reported decreased levels of pain consumed an increased dose of cannabis and/or had a different method of administration that was faster at relieving pain than that of those participants who reported no change in their pain levels. Prospective or controlled studies with a standardized product, dose, and limited route(s) of administration will be necessary to further evaluate the risks and benefits of medical cannabis for CMT.

## Conclusions

The findings from this survey indicate CMT patients report that cannabis may provide substantial symptom relief. These novel results, in conjunction with prior research, may warrant further prospective or controlled study of MC for pain management.

## Data Availability

All data produced in the present study are available upon reasonable request to the authors.

## Acknowledgments

Thanks to Iris Johnston for technical assistance. Software used in figure construction was provided by the NIEHS (T32 ES007060-31A1).

## References

[1] Stojkovic, Hereditary neuropathies: An update. Rev Neurol 2016; 172:775-778.

[2] Johnson NE, Heatwole CR, Dilik N, Sowden J, Kirk CA, Shereff D, et al. Quality of life in Charcot Marie Tooth disease: The patients perspective. Neuromuscular Dis 2014; 24: 1018–1023.

[3] Bjelica B, Peric S, Basta I, Bozovic I, Kacar A, Marjanovic A, et al. Neuropathic pain in patients with Charcot-Marie-Tooth type 1A. Neurol Sci 2020; 41(3):625–630. doi: 10.1007/s10072-019-04142-5.

[5] Carter GT, Jensen MP, Galer BS, Kraft GH, Crabtree LD, Beardsley RM, Abresch RT, Bird TD. Neuropathic pain in Charcot-Marie-Tooth disease. Arch Phys Med Rehab 1998; 79:1560–64.

[6] National Academies of Sciences, Engineering, and Medicine. 2017. The health effects of cannabis and cannabinoids: The current state of evidence and recommendations for research. Washington, DC: The National Academies Press. doi: 10.17226/24625.

[7] Basen R. New guidelines issued on medical cannabis for chronic pain— International task force shares recommendations for dosing, administering. Accessed 8/9/22 at: https://www.medpagetoday.com/meetingcoverage/painweek/88593?utm_source=Sailthru&utm_medium=email&utm_campaign=Weekly%20Review%202020-09-20&utm_term=NL_DHE_Weekly_Active

[8] Piper BJ, Dekeuster RM, Beals ML, Cobb CM, Burchman CA, Perkinson L, … Abess AT. Substitution of medical cannabis for pharmaceutical agents for pain, anxiety, and sleep. J Psychopharmacology 2017; 31(5): 569–575. DOI:10.1177/0269881117699616

[9] Hereditary Neuropathy Foundation, Accessed 8/9/2022 at: https://www.hnf-cure.org/

[10] Wallace MS, Marcotte TD, Umlauf A, Gouaux B, Atkinson JH. Efficacy of inhaled cannabis on painful diabetic neuropathy. J Pain 2015; 16:616–627.

[11] Corey-Bloom J, Wolfson T, Gamst A, Jin S, Marcotte TD, Bentley H, Gouaux B. Smoked cannabis for spasticity in multiple sclerosis: A randomized, placebo-controlled trial. CMAJ 2012; 184(10): 1143–1150. DOI:10.1503/cmaj.110837

[12] Jones E, Vlachou S. Critical review of the role of the cannabinoid compounds tetrahydrocannabinol (Δ9-THC) and cannabidiol (CBD) and their combination in multiple sclerosis treatment. Molecules 2020; 25, 4930; doi:10.3390/molecules25214930

[13] Yadav V, Bever C, Bowen J, Bowling A, Weinstock-Guttman B, Cameron M, et al. Summary of evidence-based guideline: complementary and alternative medicine in multiple sclerosis: Report of the guideline development subcommittee of the American Academy of Neurology. Neurology 2014; 82(12):1083–92. doi: 10.1212/WNL.0000000000000250.

[14] Carlini BH, Garrett SB, Carter GT. Medicinal Cannabis: A survey among health care providers in Washington State. Am J Hosp Palliat Care 2017; 34(1):85–91.DOI: 10.1177/1049909115604669

[15] Philpot LM, Ebbert JO, Hurt RT. A survey of the attitudes, beliefs and knowledge about medical cannabis among primary care providers. BMC Family Practice 2019; 20:17 https://doi.org/10.1186/s12875-019-0906-y

[16] Lombardi E, Gunter J, Tanner E. Ohio physician attitudes toward medical Cannabis and Ohio’s medical marijuana program. J Cannabis Res 2020; 2(1). DOI:10.1186/s42238-020-00025-1

[17] Braun IM,Wright A, Peteet J, Meyer FL, Yuppa DP, Bolcic-Jankovic D, et al. Medical oncologists’ beliefs, practices, and knowledge regarding marijuana used therapeutically: A nationally representative survey study. J Clin Oncol 2018; 36(19):1957–1962. doi: 10.1200/JCO.2017.76.1221.

[18] Zylla D, Steele G, Eklund J, Mettner J, Arneson J. Oncology clinicians and the Minnesota Medical Cannabis Program: A survey on medical cannabis practice patterns, barriers to enrollment, and educational needs. Cannabis Cannabinoid Res 2018; 3(1):195–202. DOI: 10.1089/can.2018.0029.

[19] Mercurio A, Aston ER, Claborn K, Waye K, Rosen RK. Marijuana as a substitute for prescription medications: A qualitative study. Subst Use Misuse. 2019; 54(11): 1894–1902. doi:10.1080/10826084.2019.1618336.

[20] Vigil JM, Stith SS, Reeve AP. Accuracy of patient opioid use reporting at the time of medical cannabis license renewal. Pain Res Manag. 2018:5704128. doi: 10.1155/2018/5704128.

[21] Bradford AC, Bradford WD. Medical cannabis laws reduce prescription medication use in Medicare part D. Health Aff 2016; 35:1230–1236. doi: 10.1377/hlthaff.2015.1661.

[22] Bradford AC, Bradford WD. Medical marijuana laws may be associated with a decline in the number of prescriptions for Medicaid enrollees. Health Aff 2017; 36(5):945–951. doi: 10.1377/hlthaff.2016.1135.

[23] Shover CL, Davis CS, Gordon SC, Humphreys K. Association between medical cannabis laws and opioid overdose mortality has reversed over time. Proc Natl Acad Sci U S A. 2019; 116(26):12624–12626. doi: 10.1073/pnas.1903434116.

[24] Kaufman DE, Nihal AM, Leppo JD, Staples KM, McCall KL, Piper BJ. Opioid mortality following implementation of medical cannabis programs in the United States. Pharmacopsychiatry 2021;54(2):91–95. doi: 10.1055/a-1353-6509.

